# Development and clinimetric testing of the Unified Drug-Induced Movement Disorders Scale (UDIMS)

**DOI:** 10.1101/2022.03.21.22272729

**Authors:** Perminder S Sachdev, Adith Mohan, Su Lynn Cheah, Anna Takacs, Jessica W Lo, Rebecca Koncz, John D Crawford

## Abstract

**Objective:** Several movement disorders develop secondary to the use of psychotropic drugs, for which multiple symptom rating scales are in common use. We planned to develop the Unified Drug-Induced Movement Scale (UDIMS) to assess the severity and impact of drug-induced dyskinesia, tremor, drug-induced parkinsonism, akathisia, dystonia and myoclonus with a single instrument.

**Methods:** Based on a literature review, consultation and pilot work, a 12-item instrument was developed, with each item rated on a 0-4 scale. The clinimetric properties of UDIMS were examined in 53 psychiatric patients on psychotropic medications, using established ratings scales for validation. The factor structure of the scale was examined, and the movement disorder correlates of distress and disability were determined.

**Results:** The instrument has good inter-rater reliability. Its correspondence with three other scales – Abnormal Involuntary Movements Scale, Simpson-Angus Scale and Prince Henry Hospital Akathisia Scale – for the relevant items was high. A principal components analysis yielded four factors, considered to represent tremor, parkinsonism, akathisia and dyskinesia. Overall movement-disorder related disability was related to parkinsonism and dyskinesia, while distress to all four components.

**Conclusions:** UDIMS is a reliable and valid scale to quantify a range of drug-induced movement disorders (DIMDs), that obviates the need for the use of multiple rating scales. Its widespread use by both clinicians and researchers, and further refinement based on this, will help promote the detection and treatment of drug-induced movement disorders, thereby reducing both distress and disability.

## INTRODUCTION

Several movement disorders are known to develop in association with psychotropic drugs. While first- and second-generation antipsychotics (FGAs and SGAs) are the most prominent causes of drug-induced movement disorders (DIMDs) (1), they also develop with most other classes of psychotropic drugs including, though not limited to, antidepressants (tricyclics or TCAs and selective serotonin reuptake inhibitors or SSRIs), lithium, stimulants, antiepileptic mood stabilizers such as sodium valproate and carbamazepine, and anticholinergic drugs (2).

The manifestations of DIMDs vary widely, and include dyskinesia (especially tardive), tremor, drug-induced parkinsonism, akathisia, dystonia and myoclonus. It is well established that these motor symptoms can be disabling and distressing for patients, and negatively impact their quality of life (3). They can be a significant disincentive to adherence with prescribed medication, hence contributing to illness relapses, and adverse clinical and socio-vocational outcomes. Screening for and systematic monitoring of DIMDs is therefore essential for safe and effective clinical care. Equally, in research settings, the reliable assessment of DIMD severity is necessary for the investigation of side-effect profiles of various drugs, the study of relationships between motor disorders and other clinical variables, and in advancing our understanding of the neuropathological underpinnings of DIMDs such as tardive dyskinesia (TD), so that treatments can be developed.

Although several reliable and valid symptom rating scales are in common usage, these have been designed to assess specific symptom domains rather than the entire range of DIMDs. Examples include the Simpson-Angus Scale for drug-induced parkinsonism (SAS) (4), the Prince Henry Hospital Akathisia Rating Scale for akathisia (PHHARS) (5), and the 12-item Abnormal Involuntary Movements Scale (AIMS) (6) for the assessment and monitoring of TD. Psychiatric patients are often likely to be treated with multiple psychotropics, and each drug is in turn associated with several movement disorders. In a patient who presents with a combination of motor symptoms, it is then necessary to use multiple rating scales at each review, thereby presenting a challenge to the time-poor clinician or researcher. Hence there is a need for a combined rating scale that obviates the requirement for multiple scales and allows for a more global assessment of the patient with a DIMD, while addressing the heterogeneity which is common in such presentations.

We present the development and clinimetric evaluation of the Unified Drug-Induced Movement Scale (UDIMS). It is a 12-item clinician-rated scale designed to assess the severity and impact of dyskinesia, tremor, drug-induced parkinsonism, akathisia, dystonia and myoclonus due to psychotropic drugs, developed as a simple to administer tool for use in adults.

## METHOD

### Scale development

The authors reviewed the literature and consulted informally with other clinicians before including the following movement disorders in the scale: dyskinesia, akathisia, drug-induced parkinsonism, tremor, dystonia and myoclonus (2). Neuroleptic malignant syndrome (NMS) was not included because of the complexity of the syndrome, with movement disorder (rigidity and/or catatonia) being only one of several salient features (7). Catatonia is also a complex and uncommon syndrome that warrants its separate clinical evaluation (8). It was decided to base the rating of the movement disorder on clinical observation alone, and not take etiology into consideration, as the latter is a clinical determination based on historical and other data. The rating of dyskinesia does not therefore consider whether the onset is acute or tardive. The rating of action (or postural) tremor, as for other movements, is irrespective of which drug is causative, or if multiple drugs are contributory. The ratings reflect observations by the clinician cross-sectionally only.

Since dyskinesia can manifest in several body regions, with nine body regions being rated by AIMS, a decision was made to reduce these to two clusters – orolingual-buccal-facial (OLBF) and limb-truncal (LT) – based on prior evidence that these regions are known to cluster, which may have treatment and prognostic implications (9, 10). The rating scale of 0- 4 used in AIMS was considered the most appropriate, with a rating of ‘1’ for minimal (or uncertain) presence and ‘0’ representing a definite absence. A pilot joint evaluation of 10 cases of TD was used to decide on the method of rating mild ‘2’, to severe ‘4’ in each of the clusters.

Tremors seen in psychiatric patients are frequently present on action (posture holding or kinetic) or at rest (parkinsonian) (11). The other parkinsonian features of note are rigidity and bradykinesia (12). Akathisia has two components – subjective and objective – and since either can be present without the other (5), both items were chosen to be rated. Dystonia and myoclonus are other frequent manifestations related to medications (2).

For uniformity of scoring, all items are rated from 0 to 4 as described above. In the pilot phase, guidelines were developed for the rating of these items (Appendix 1). Subtotals are calculated for the items grouped together (e.g. dyskinesia, akathisia, etc.) and a grand total provides an overall severity measure, while acknowledging the heterogeneity. There is also a global rating of distress and disability, similar to what is done in AIMS. Disability refers to the functional impairment produced by the movement disorder, whereas distress is subjective and may be due to social embarrassment, interference with function or pain. A particular movement, such as orolingual dyskinesia, may be very distressing but may not impair the patient’s functioning. On the other hand, severe dystonia can be both severely distressing and disabling.

### Clinimetric evaluation

<u>Sample</u>: The sample comprised 53 adult (>18 years) patients presenting to the Eastern Suburbs Mental Health Services of South-Eastern Sydney Local Health District. Participants were included in the study if they suffered from a major psychiatric disorder, were currently being treated with one or more psychotropic drugs, had received the medication for 3 months or more, had sufficient proficiency in English to complete the interview and rating scales, and were able to provide written informed consent to participate. The presence of substance use disorder or a primary neurocognitive or neurodevelopmental disorder was exclusionary.

The study was approved by the Human Research Ethics Committee of the South-Eastern Sydney Local Health District, and all participants provided informed consent.

#### Procedure

The participants were interviewed by one of the investigators for sociodemographic data, a brief structured medical and psychiatric history, medication history and list of current medications. The UDIMS was completed with a standardized procedure, as described in Appendix 1. In addition, the following scales were administered: Abnormal Involuntary Movements Scale (AIMS), Simpson Angus scale (SAS), and Prince Henry Hospital Akathisia Rating Scale (PHHRS). A proportion (about 1 in 5) of assessments were videotaped, with consent, for independent rating by two other raters.

### Statistical analysis

Summary statistics of age and sex were calculated. To compute the proportion of participants with each movement disorder, UDIMS scores and a cut-off of 2 and above were used, with a score of 2 indicating definite presence of a movement abnormality.

First, we examined how UDIMS corresponds to the established scales (AIMS, SAS or PHAS). Table 1 shows how each UDIMS item corresponds to an item or a combination of items from an established scale. Raw scores were transformed into z-scores and for when there were multiple items for the corresponding scales, we calculated the sum of the z-scores (the composite). Since most scores were skewed and had less than 4 categories, the Spearman’s rank-order correlation coefficient between an UDMIS item and a corresponding composite was calculated. To obtain the 95% confidence interval (95% CI) of the Spearman’s coefficients, scores were transformed to the rank order distribution to obtain the 95% CI of the Pearson correlations (which correspond to the Spearman’s). In addition, the weighted Kappa statistic between each UDIMS item and a corresponding scale item(s) was calculated using raw scores or mean raw scores rounded to the nearest integer. For UDIMS items 7 and 8, the rating items 1 and 2 were combined based on clinical decision to form a 4-point scale to match with the PHAS scale. The weighting matrix used for the 5-point and 4-point scales were 1 \ 0.67 1 \ 0.33 0.67 1 \ 0 0.33 0.67 1 and 1 \ 0.75 1 \ 0.5 0.75 1 \ 0.25 0.5 0.75 1 \ 0 0.25 0.5 0.75 1 respectively.

**Table 1:** Correlation and agreement between each UDIMS item and corresponding composite scores from comparison scales [AIMS: Abnormal Involuntary Movement Scale (6); SAS: Simpson and Angus Scale (4); PHAS: Prince Henry Akathisia Scales (5)]

| UDIMS item | Movement disorder | Corresponding scale | Spearman coefficient (95% CI; p-value) | Kappa (weighted)<sup>a</sup> |
| --- | --- | --- | --- | --- |
| 1 | Dyskinesia – oro-lingual-buccal-facial | AIMS I (1, 2, 3, 4) | 0.87<br>(0.79, 0.93; <0.001) | 0.68 |
| 2 | Dyskinesia – limbs & trunk | AIMS II and III (5, 6, 7) | 0.68<br>(0.50, 0.80; <0.001) | 0.69 |
| 3 | Tremor – at rest | SAS 9 | 0.86<br>(0.77, 0.92; <0.001) | 0.72 |
| 4 | Tremor – on action | SAS 9 | 0.69<br>(0.52, 0.81; <0.001) | 0.60 |
| 3 & 4 | Tremor combined | SAS 9 | 0.88<br>(0.80; 0.93; <0.001) | 0.71 |
| 5 | Muscle rigidity | SAS 2, 3, 4, 5, 6 | 0.86<br>(0.77, 0.92; <0.001) | 0.39 |
| 6 | Bradykinesia | No scale | NA | NA |
| 7 | Akathisia subjective | PHAS Subjective 1, 2, 3 | 0.71<br>(0.54, 0.82; <0.001) | 0.61 |
| 8 | Akathisia objective | PHAS Objective I 1, 2, 3, 4; II 1, 2, 3 | 0.74<br>(0.59, 0.85; <0.001) | 0.25<sup>b</sup> |
| 9 | Dystonia | No scale | NA |  |
| 10 | Myoclonus | No scale | NA |  |
| 11 | Total disability | No correlate | NA |  |
| 12 | Total distress | No correlate | NA |  |

<sup>a</sup> Based on mean raw scores, rounded to the nearest integer. For UDIMS items 7 and 8, the rating 1 and 2 were combined to form a 4-point scale to match with the PHAS scale.
<sup>b</sup> All patients have low scores for PHAS objective items. Combined scores rounded to the nearest integer are 0 and 1 only (48 zeros and 3 ones).

Regression was used to examine how UDIMS items predicted disability (UDIMS 11) and distress (UDIMS 12). Since scores were skewed, generalized linear model with Poisson distribution and log link was used. For variable selection, first, a backwards stepwise method based on p<0.1 was used. Then the Akaike information criterion (AIC) or the likelihood-ratio test (when a model is nested within another, and the sample size is the same) were used to find a model with best fit. UDIMS raw scores rather than standardized scores were used to aid interpretation. We conducted unadjusted and adjusted (by sex, age, primary psychiatric diagnoses) analyses.

Principal component analysis (PCA), with oblique rotation to simple structure using the Oblimin procedure, was used to examine the correlational structure of ratings on the UDIMS items. The number of components extracted was determined by examination of the values of component eigenvalues in the unrotated solution and the interpretability of the solutions. Solutions with component eigenvalues greater than one, or slightly less than one, were examined and used for further statistical analyses, depending on the meaningfulness of the solution. Composite UDIMS subscales were formed by combining UDIMS items loadings on the same components in the rotated PCA solutions. The subscales were calculated as the average of the z-scores of the component items and used in a generalized linear model (Poisson family with log link) to examine patterns of comorbidity. Subscales with p<0.1 were retained in the model. UDIMS item 10 – myoclonus was not included in the regression and the PCA since no patients had this movement disorder; UDIMS item 9 was also not included since only one patient had dystonia.

Two authors, both neuropsychiatrists (PSS and AM), completed the UDIMS scale on 10 patients based on video recordings to assess the scale’s inter-rater reliability. We calculated Gwet’s AC1 to assess the degree of agreement between three raters. We used a weighting matrix of (1 \ 0.8 1 \ 0.5 0.8 1 \ 0.2 0.5 0.8 1; Table S1) to describe the degree of agreement between raters for different categories of the scale.

All statistical analyses were performed using Stata 15.1 and IBM SPSS Statistics 26.0.

## RESULTS

### Description of the sample

The mean age of the sample (N=53) was 44.8 years (SD 13.0, range 20-67), with 28 (53%) men. The main psychiatric diagnoses were schizophrenia (47%), bipolar disorder (15%), major depression (15%), schizoaffective disorder (11%) and other (12%). The movement disorders represented were dyskinesia (n=10; 19%), tremor (n=12; 23%), parkinsonism (n=7; 13%), akathisia (n=9; 17%) and dystonia (n=1; 1.9%). No participants had myoclonus.

#### Reliability

The weighted Gwet’s AC1 for items 1-8 ranged from 0.89 to 0.99 (all p<0.001); item 10 had full agreement between raters (Table S2). The global severity rating (item 11) had a reliability score of 0.89 (95%CI 0.68-1.00, p<0.001), and global distress 0.76 (95%CI 0.55-1.00, p<0.001). The details are presented in Table 3S.

#### Validity

The correlations between UDIMS items and the corresponding measures of the comparison scales based on standardize z-scores were high, Spearman’s ρ ranging from 0.69 to 0.92 (p<0.001 for all) and weighted kappa coefficients 0.25 to 0.72 (Table 1). The dystonia and myoclonus items could not be examined because of their infrequency. The correlations with non-corresponding items were uniformly low except for UDIMS item 7 with objective akathisia on PHAS (ρ = 0.58) and UDIMS item 8 with subjective akathisia on PHAS (ρ = 0.57), reflecting the correlation between the two features of akathisia. The total disability and distress scores did not have any corresponding ratings for comparison.

### Factor structure

The results of the principal component analyses are shown in Tables 2 and 3. The rotated component solutions can clearly be interpreted based on the classification of UDIMS items shown in Appendix 1. Root-one criteria (i.e., all component eigenvalues greater than one) produced a 3-component solution which is shown in Table 2. Components PC2_3_ and PC3_3_ are defined by pairs of items which are indicators of dyskinesia and akathisia, respectively. Component PC1_3_ is defined by a combination of 2 pairs of items, 2 listed in Appendix 1 as indicators of Tremor and 2 of parkinsonism. In light of this finding, a 4-component solution was examined in order to determine whether this would achieve a separation of the two sets of items defining PC1_3_. The 4-component solution, shown in Table 3, reveals that this was achieved. Here the 4 components, PC1_4_ to PC4_4_ are each clearly defined by pairs of items, listed in Appendix 1 as indicators of tremor, dyskinesia, akathisia and parkinsonism, respectively. These findings can be interpreted as an empirical verification of the validity of the classification of items as shown in Appendix 1.

**Table 2:** Principal component analysis with oblique rotation to simple structure: 3-factor solution

| **Pattern Matrix** |  |  |  |  |
| --- | --- | --- | --- | --- |
| **UDIMS items** | **PC 1<sub>3</sub>** | **PC 2<sub>3</sub>** | **PC 3<sub>3</sub>** | **h<sup>2</sup>** |
| 1 Dyskinesia – oro-lingual-buccal-facial | -0.14 | 0.85 | 0.02 | 0.72 |
| 2 Dyskinesia – limbs & trunk | 0.07 | 0.88 | 0.03 | 0.79 |
| 3 Tremor – at rest | 0.82 | -0.18 | 0.06 | 0.69 |
| 4 Tremor – on action | 0.83 | -0.16 | 0.20 | 0.77 |
| 5 Muscle rigidity | 0.57 | 0.11 | -0.40 | 0.46 |
| 6 Bradykinesia | 0.69 | 0.18 | 0.10 | 0.55 |
| 7 Akathisia subjective | -0.02 | -0.09 | 0.88 | 0.78 |
| 8 Akathisia objective | 0.26 | 0.22 | 0.79 | 0.78 |
| Eigenvalues of unrotated solution | 2.49 | 1.64 | 1.40 |  |

| **Component Correlation Matrix** |  |  |  |
| --- | --- | --- | --- |
|  | **PC 1<sub>3</sub>** | **PC 2<sub>3</sub>** | **PC 3<sub>3</sub>** |
| **PC1<sub>3</sub>** | 1.00 | 0.07 | 0.11 |
| **PC2<sub>3</sub>** | 0.07 | 1.00 | -0.04 |
| **PC3<sub>3</sub>** | 0.11 | -0.04 | 1.00 |

PC = principal component ; h<sup>2</sup> = communalities

**Table 3:**
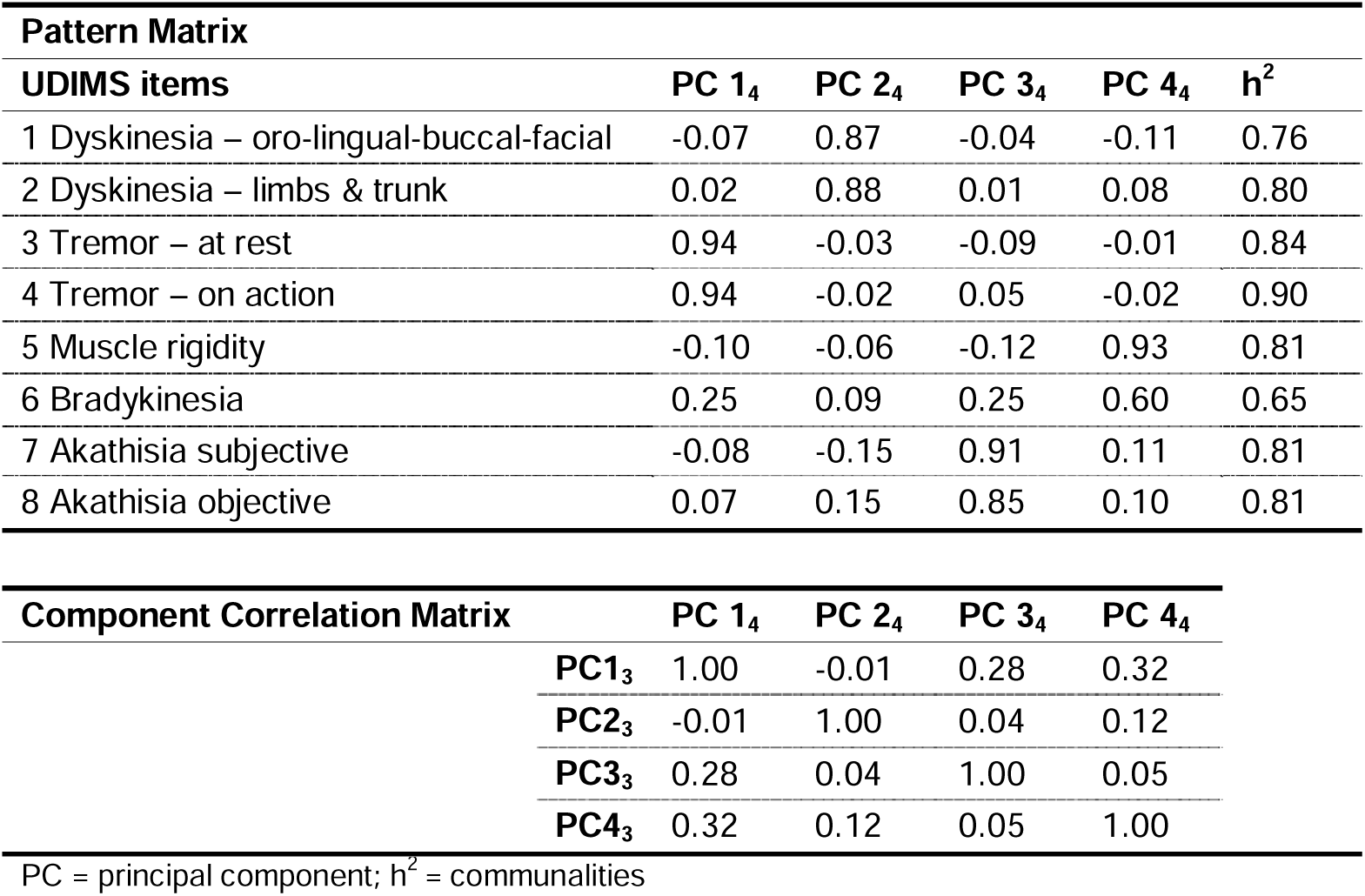
Principal component analysis with oblique rotation to simple structure: 4-factor solution

#### Distress and Disability

To examine the determinants of disability and distress, a regression analysis was performed, adjusting for age, sex and psychiatric diagnosis (Table 4). Disability on UDIMS was best related to items 1 (oro-lingual-buccal-facial dyskinesia), 3 (tremor at rest) and 6 (bradykinesia). Distress was related to items 2 (limb-truncal dyskinesia), 3 (tremor at rest) and 7 (subjective akathisia). When the subscales formed from the components of the PCA were used in the regression analysis, disability was related to parkinsonism and dyskinesia, while distress to all three components.

**Table 4:**
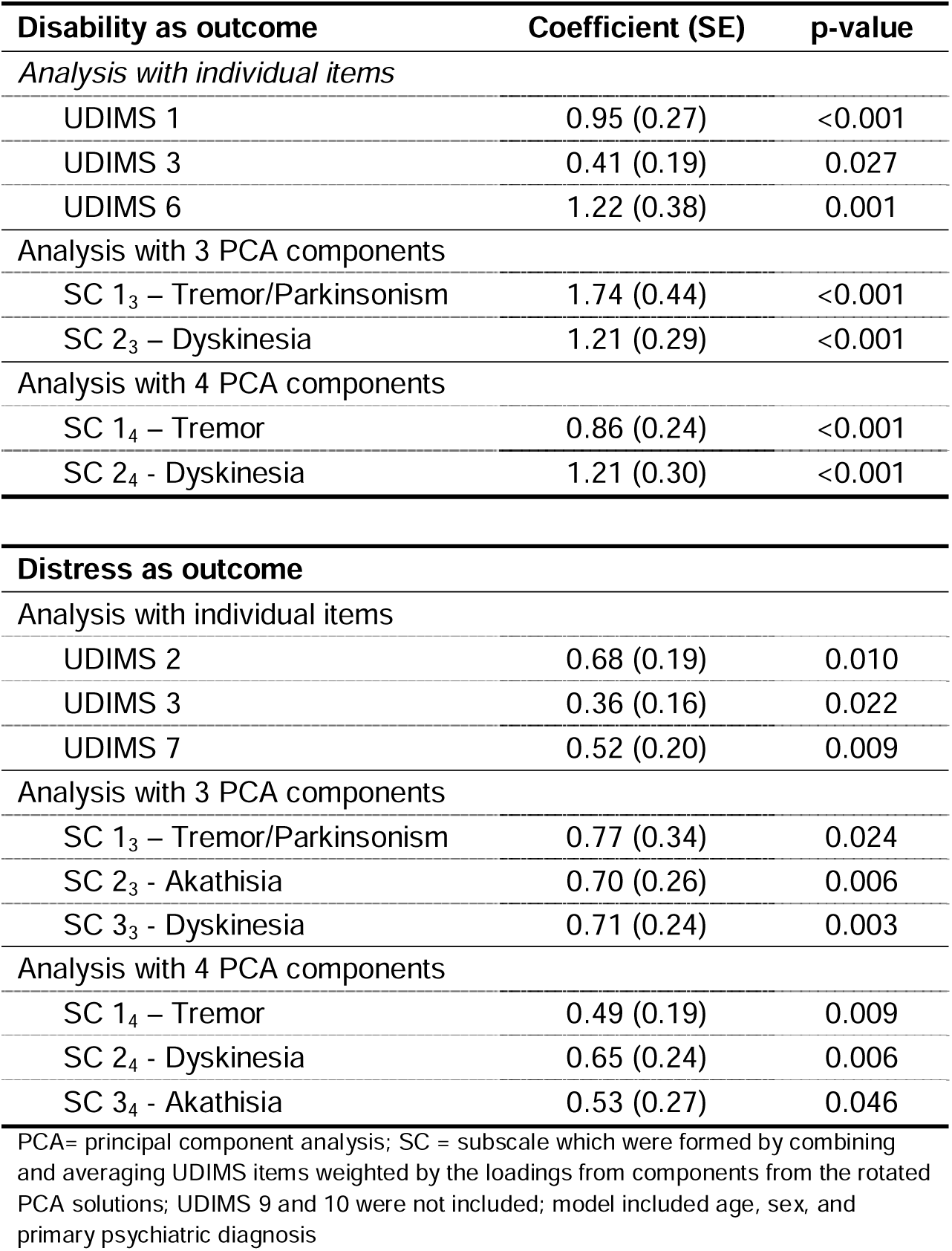
Regression analysis exploring predictors of disability (UDIMS11) and distress (UDIMS 12)

| **Disability as outcome** | **Coefficient (SE)** | **p-value** |
| --- | --- | --- |
| *Analysis with individual items* |  |  |
| UDIMS 1 | 0.95 (0.27) | <0.001 |
| UDIMS 3 | 0.41 (0.19) | 0.027 |
| UDIMS 6 | 1.22 (0.38) | 0.001 |
| *Analysis with 3 PCA components* |  |  |
| SC 1<sub>3</sub> – Tremor/Parkinsonism | 1.74 (0.44) | <0.001 |
| SC 2<sub>3</sub> – Dyskinesia | 1.21 (0.29) | <0.001 |
| *Analysis with 4 PCA components* |  |  |
| SC 1<sub>4</sub> – Tremor | 0.86 (0.24) | <0.001 |
| SC 2<sub>4</sub> - Dyskinesia | 1.21 (0.30) | <0.001 |
| **Distress as outcome** |  |  |
| *Analysis with individual items* |  |  |
| UDIMS 2 | 0.68 (0.19) | 0.010 |
| UDIMS 3 | 0.36 (0.16) | 0.022 |
| UDIMS 7 | 0.52 (0.20) | 0.009 |
| *Analysis with 3 PCA components* |  |  |
| SC 1<sub>3</sub> – Tremor/Parkinsonism | 0.77 (0.34) | 0.024 |
| SC 2<sub>3</sub> - Akathisia | 0.70 (0.26) | 0.006 |
| SC 3<sub>3</sub> - Dyskinesia | 0.71 (0.24) | 0.003 |
| *Analysis with 4 PCA components* |  |  |
| SC 1<sub>4</sub> – Tremor | 0.49 (0.19) | 0.009 |
| SC 2<sub>4</sub> - Dyskinesia | 0.65 (0.24) | 0.006 |
| SC 3<sub>4</sub> - Akathisia | 0.53 (0.27) | 0.046 |

PCA= principal component analysis; SC = subscale which were formed by combining and averaging UDIMs items weighted by the loadings from components from the rotated PCA solutions; UDIMs 9 and 10 were not included; model included age, sex, and primary psychiatric diagnosis

#### User experience

The scale takes 7-10 minutes to administer. This duration can be reduced if it is administered as part of a full psychiatric evaluation of a patient. It has been so far administered by psychiatrists and trainee psychiatrists with previous experience of movement disorders in the setting of psychiatric illness. It takes 3-5 sessions to achieve good inter-rater reliability in its administration.

## DISCUSSION

UDIMS was developed to provide a reliable and valid assessment of the full range of drug-induced movement disorders in psychiatric patients. The intention was to obviate the need to use several rating scales designed specifically to rate individual disorders such as dyskinesia, akathisia, parkinsonism, tremor, etc., making it easier for a clinician or researcher to document the severity of the full range of movement disorders in their patients. UDIMS uses a rating strategy that is well-established in the field, since the advent of the AIMS in 1976 (6). It is therefore not surprising that it achieved good inter-rater reliability and its correspondence with the individual scales was high. It also has good intra-rater reliability, and new raters with experience in the treatment of psychiatric disorders can be easily trained to administer it reliably.

UDIMS is a composite scale with items that can be aggregated to provide subscales, specifically for tremor, dyskinesia, parkinsonism and akathisia. The PCA showed a good separation of these subscales, with the correlations between items being high within each subscale, but low with other items without. This is unsurprising as these DIMDs do not generally co-occur, manifesting at different times in the treatment trajectory and differentially with various drugs. The exception is the common co-occurrence of acute akathisia and drug-induced parkinsonism (13), since both occur early in the course of antipsychotic treatment. The correlation of the objective akathisia item with the sum of the parkinsonism items was 0.35, which is consistent with the correlation of 0.3 between akathisia and parkinsonism ratings reported previously (13).

Special comments are warranted in relation to tremor, myoclonus and dystonia. Tremor is a common side effect of psychotropic medication, occurring either *de novo* or as an exaggeration of an underlying tremor related to the medical and psychiatric conditions of the patient (11). Tremors are often classified according to their activation conditions, occurring either at rest or upon action, usually postural or kinetic (14). Resting tremors have been typically associated with antipsychotic drugs, whereas action tremors occur with a range of psychotropics, including antidepressants and antiepileptics. Lithium has been associated with both resting and action tremor, and antipsychotics not uncommonly also result in action tremor (11). The correlation between the two types of tremor was high in our sample, partly reflecting the fact that the patients were being treated with multiple medications. For the drug-induced parkinsonism subscale, we recommend a summation of resting tremor, rigidity and bradykinesia, and to not include action tremor even though it loaded on PC1 in the 3-factor solution. We could not analyze myoclonus and dystonia in our sample. Myoclonus, while not uncommon with some psychotropics such as the SSRIs (15), is often difficult to capture in a brief rating scale because of its intermittent occurrence. Acute dystonia, the more common form of drug-induced dystonia (16), is unlikely to occur in the sample included in this study, and tardive dystonia has a low prevalence (17), such that only one patient in this sample had the disorder. The application of the scale to larger samples will help determine the reliability of assessing these two DIMDs.

Movement disorders produce distress and disability, but the relative contributions of the various DIMDs differ. While parkinsonism and dyskinesia were noted to contribute to both disability and distress, akathisia contributed significantly to distress and not disability. The inclusion of these two items to the scale adds measures of the total impact of the DIMDs, which the sum total of the individual items does not capture effectively. In our study, the correlations of distress and disability with total score (sum of UDIMS items 1 to 10) were 0.67 (p<0.001) and 0.70 (p<0.001) respectively, and the correlation between themselves was 0.54 (p<0.001), highlighting the utility of both items in completing the clinical picture. This scale has been developed as a tool for the documentation of the severity of DIMDs, and is not designed to be a diagnostic assessment, the latter being a clinical determination. While the dyskinesia being rated is likely to be due to TD, the diagnosis of TD is independent of the rating and is based on a full clinical evaluation of the patient. In the same vein, the etiology of tremor requires a thorough evaluation of the patient. Moreover, the rating scale is a cross-sectional assessment and longitudinal evaluation is necessary for several clinical decisions. Repeated administration may assist this process, and video-recording of the UDIMS interview may be considered for this, after appropriate patient consent.

We acknowledge a number of limitations of UDIMS. First, in the attempt to capture a wide range of DIMDs, compromise on detailed evaluation had to be accepted. For instance, while AIMS has 7 items to rate dyskinesias in various body regions, this was collapsed to 2 items in UDIMS, thereby losing detail. It is arguable that the more detailed evaluation may be necessary for certain purposes, but the trade-off was necessary to make the instrument practical. Second, there is inherent heterogeneity in the scale owing to the designed capture of several DIMDs that do not necessarily co-occur. The scale therefore lends itself to subscales, particularly those for dyskinesia, parkinsonism and akathisia, as well as independent ratings of tremor, dystonia and myoclonus, and should be regarded as a composite scale of related disorders. Third, the total score is a sum of the various subscales, rather than an overall rating given by the examiner. This is deliberate, since pilot work demonstrated a difficulty in reliably assigning a global severity score when a range of disorders had to be considered. On the other hand, global distress and disability, as indicators of global severity, were more reliably rated. Fourth, our sample is relatively small, and while adequate to assess the clinimetric properties of most items, we could not determine the reliability of the ratings of myoclonus and dystonia. Much larger samples are needed for this purpose, and we hope that the widespread use of this instrument will lead to its refinement and improvement, as has happened for other instruments such as the MDS-Unified Parkinson’s Disease Rating Scale (MDS-UPDRS) (18). Fifth, its reliability has been examined by psychiatrists and trainees with considerable experience in treating major psychiatric disorders with drugs. Its performance in the hands of other professionals without such experience remains to be determined. Sixth, even though intending to cover the full range of DIMDs, UDIMS does not include catatonia and neuroleptic malignant syndrome (NMS). This is because these are complex syndromes with movement disorder being but one of the salient features of each. Other scales specifically designed for catatonia (8) and NMS (19) may be more appropriate for these syndromes.

## CONCLUSION

We present UDIMS as a reliable and valid scale to quantify a range of DIMDs, to be applied by both clinicians and researchers. With its widespread use, and further refinement based on this, it will obviate the need for the use of multiple rating scales to capture the full range of DIMDs, and promote the detection and treatment DIMDs in psychiatric patients, thereby reducing both distress and disability.

## Data Availability

All data produced in the present work are contained in the manuscript

## Acknowledgements

We thank the referring clinicians and patients of the Eastern Suburbs Mental Program of the South Eastern Sydney Local Health District.

## SUPPLEMENTARY MATERIAL

**Table S1:** Weighting matrix used for the inter-rater reliability analysis

|  | 0 – normal | 1 – minimal /<br>borderline | 2 – mild | 3 – moderate | 4 – severe |
| --- | --- | --- | --- | --- | --- |
| 0 – normal | 1 | 0.8 | 0.5 | 0.2 | 0 |
| 1 – minimal / borderline | 0.8 | 1 | 0.8 | 0.5 | 0.2 |
| 2 – mild | 0.5 | 0.2 | 1 | 0.8 | 0.5 |
| 3 – moderate | 0.2 | 0.5 | 0.8 | 1 | 0.8 |
| 4 – severe | 0 | 0.2 | 0.5 | 0.8 | 1 |

**Table S2.** Inter-rater reliability analysis: weighted Gwet’s AC1

| **UDIMS items** | **Weighted Gwet's AC1** | **95% CI; p-value** |
| --- | --- | --- |
| UDIMS 1 | 0.89 | 0.64, 1.00; <0.001 |
| UDIMS 2 | 0.93 | 0.73, 1.00; <0.001 |
| UDIMS 3 | 0.99 | 0.95, 1.00; <0.001 |
| UDIMS 4 | 0.97 | 0.92, 1.00; <0.001 |
| UDIMS 5 | 0.94 | 0.86, 1.00; <0.001 |
| UDIMS 6 | 0.97 | 0.92, 1.00; <0.001 |
| UDIMS 7 | 0.94 | 0.67, 1.00; <0.001 |
| UDIMS 8 | 0.97 | 0.92, 1.00; <0.001 |
| UDIMS 9 | NA (perfect match) | NA |
| UDIMS 10 | 0.99 | 0.95, 1.00; <0.001 |
| UDIMS 11 | 0.89 | 0.68, 1.00; <0.001 |
| UDIMS 12 | 0.76 | 0.55, 1.00; <0.001 |

## APPENDIX 1

### UNIFIED DRUG-INDUCED MOVEMENT DISORDERS SCALE (UDIMS)

#### Description

The UDIMS is a 12-item clinician-related scale designed to assess the severity and impact of movement disorders secondary to psychotropic drug use, in particular due to antipsychotic drugs. It has items to assess dyskinesia (especially tardive), tremor, drug-induced parkinsonism, akathisia, dystonia and myoclonus. It is simple to administer and does not require any specific instruments. It has been developed for use in adults. It obviates the need for multiple rating scales for the various drug-induced movement disorders (DIMD).

The scale is not a diagnostic instrument, and not a substitute for clinical judgement in the diagnostic assessment. The establishment of a drug-related etiology is a clinical determination. The scale is intended to document the severity of the movement disorder in the context of psychotropic use. While it is most likely to be useful in assessing patients being treated with antipsychotic drugs, it is also applicable for patients on antidepressants, mood-stabilizing drugs such as lithium carbonate and sodium valproate, and other such drugs, all of which may be associated with movement disorders.

#### Scoring

Each item is scored on a ‘0’ (normal), ‘1’ (slight, borderline abnormal), ‘2’ (mild), ‘3’ (moderate) and ‘4’ (severe) scale. A score of ‘2’ or higher indicates a definite abnormality. A score of 1 raises the suspicion of an abnormality worthy of follow-up. There is an item each for the disability and distress produced by the DIMDs.

#### Examination procedure

1. The examination process begins with unobtrusive observation of the patient in the interview room and continues through the interview process. If this rating is part of a longer interview, it is best to conduct the ratings towards the end of the interview to allow more observation time.
2. The patient should be examined in both the sitting and standing positions. Most of the interview is best conducted with the patient sitting in a straight back chair without arms. The patient’s limbs should be partially uncovered, and all clothes (e.g. coats, shawls etc.) that might block the visualization of movements should be removed. Make certain that the full profile of the patient is visible to you.
3. The patient should not be chewing gum or candy. He/she should not be holding anything in the hands. Ask if patient is wearing dentures, and if there is pain or discomfort in the teeth.
4. Observe the patient as you converse with him/her for a few minutes. Pay attention to all parts of the body for any abnormal movements.
5. Ask the patient to sit with arms resting on the legs so that the hands are unsupported. Observe for tremor in the hands at rest. Also look for movements in other parts of the body.
6. Ask patient to extend their arms in front of them, parallel to the ground and with palms facing down and fingers slightly spread out. Observe for action tremor. Observe for dyskinetic movements in face, trunk, and legs as well [due to activation].
7. Ask the patient to open their mouth and keep it open while you observe for any choreoathetoid movement in the tongue as it rests in the mouth. Repeat it once. Then ask the patient to protrude their tongue and observe for abnormal movements in the tongue, perioral region as well as the rest of the body [activation].
8. Ask patient to tap thumb to each finger for 10-15 sec, first with one hand and then the other [another activation maneuver]. Observe for activation of dyskinetic movements in all parts of the body.
9. Ask the patient if they have feelings of restlessness, and whether they have difficulty in sitting or standing in one place to any length of time. Ask if they have difficulty in keeping their legs still when sitting or standing. Subtle abnormalities are noticeable then the individual is in a queue at a supermarket or stands to cook a meal. Ask if the restlessness is in the body or the mind or both, and whether there is a clear relationship with the initiation of medication or the time of day when the drug is taken.
10. Rigidity is best examined at the elbow and wrist. With the elbow bent at right angles, passively extend and flex the elbow joint, with the patient relaxed and then asked to move their head from side to side [activation]. Then test muscle tone at the wrist, with the wrist in one hand the fingers held in the examiner’s other hand as the wrist moved to extension, flexion, radial and ulnar deviation. Repeat with activation. Do on both sides. Ignore cogwheeling when assessing rigidity.
11. Look at patient’s facial expression and any changes through the interview. Look for slowness of movement, salivation, and poverty of gestures. Ask patient to open and close hand in rapid succession, first one and then the other.
12. Ask patient to extend both arms with palms facing down for 10 sec. Then, ask patient to extend wrists for 10 secs. Then, patient performs finger to nose movement four times, first with one hand and then the other. Observe for myoclonus and tremor.
13. Ask patient to fold both arms across the chest and get up from the sitting position to stand up. If unable, examiner may help the patient stand up.
14. Patient stands with feet as close together as is comfortable as examiner talks to them for 30-60 secs. Observe the patient’s movements in this period.
15. Ask patient to extend their arms in front of them for 10 sec. as before.
16. Ask patient to turn around and walk 6 m at their usual speed, turn around and come back. Observe for arm swing, gait speed, festination, poor balance, or abnormal movements.
17. Ask patient to write their name and draw a spiral with one hand and then the other.

Please complete the examination procedure and rate the movements as present during the examination period. The ratings are based on the overall observations.

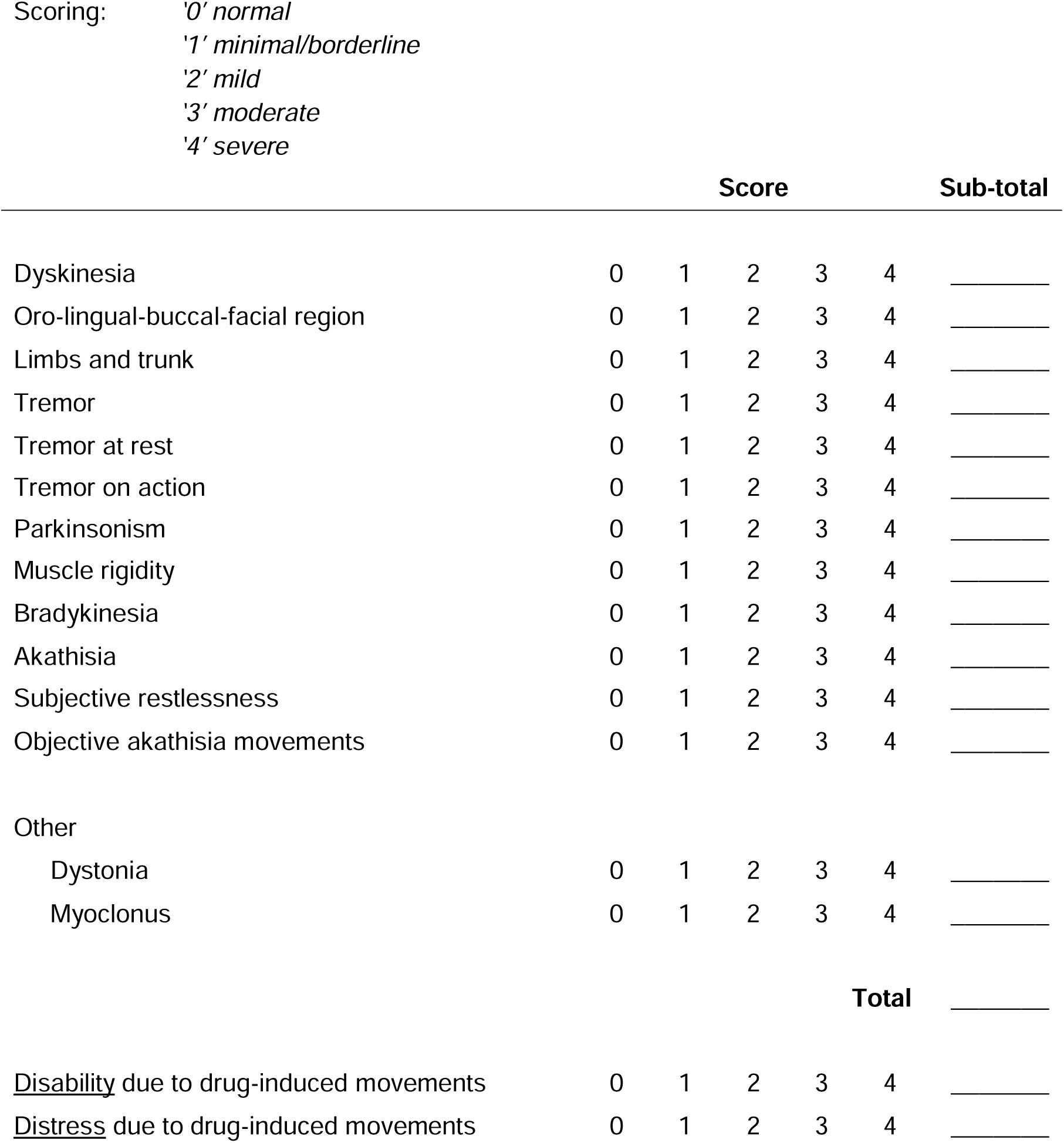

### SCORING PROCEDURE – SOME GUIDELINES

#### Dyskinesia

The ratings are for choreoathetoid movements of the oro-lingual, buccal, facial as well as limb-truncal regions. The movements include blinking, facial grimacing, puckering, lip smacking, jaw clenching, chewing, mouth opening, tongue protrusion, etc. The limb movements may be rapid, purposeless, and irregular, or slow and irregular. Truncal movements include rocking, twisting, squirming, diaphragmatic movements, and pelvic movements. The earlier movements are often tongue movements with the tongue sitting at the base of the mouth. Activation maneuvers tend to exacerbate dyskinetic movements or bring them out. If present only during activation, rate them lower.

#### Tremor

Tremor is usually most obvious in the fingers and hands although it may occur in upper and lower extremities and the head. Tremor affecting multiple body regions is more severe that that affecting one body region only. Parkinsonian tremor is generally present at rest. Action tremor is associated with several psychotropic drugs including lithium, antidepressants, valproate, etc. The rating of the severity of the tremor is based on both the amplitude of the tremor and how persistent it is during the examination. A rating of 1 is given to a tremor that is slight and infrequently present or may be a slight exaggeration of the physiological tremor. Such tremors may only be apparent when the individual is very anxious. A rating of mild ‘2’ is for tremors that are mild in amplitude but persistent, or moderate but only infrequently present. Severe ‘4’ tremors are present for most of the time and for action or postural tremors, interfere with activities such as feeding, dressing, etc. Writing or drawing are useful to document the amplitude and frequency of a tremor.

#### Parkinsonism

The rating of rigidity is an overall rating, based on the testing of upper and lower limb muscles, although upper limb muscles of the wrist and elbow are the ones usually examined. Slight increase in muscle tone may be apparent only on activation by movement of other muscles such as turning the head. In moderate rigidity, full range of passive movement is still achievable without difficulty, but in severe rigidity, this poses great difficulty. Bradykinesia is a combination of slowness, hesitancy, smaller amplitude of movements, reduced responsiveness and poverty of spontaneous movements. In minimal ‘1’ cases, the individual may appear normal to those not familiar with his/her premorbid state. In mild cases, there is slowness which is definitely abnormal, and the amplitude of movements may be reduced. This may be associated with reduced arm swing while walking. While drooling may be associated with bradykinesia, it can also be due to increased salivation without bradykinesia and this should be considered.

#### Akathisia

There are two aspects to akathisia, a subjective component and the presence of movements that can be voluntarily suppressed for varying periods of time. Each is rated separately. Minimal ‘1’ rating for a non-specific feeling of restlessness, which may be difficult to distinguish from anxiety or agitation. For mild ‘2’ rating, the patient has a definite awareness of inner restlessness, often referred to the legs, which is aggravated by the requirement to keep still, but this is mild and does not interfere with functioning. In severe ‘4’ cases, the restlessness is unrelenting and is a major problem for the individual.

For objective rating, the patient is observed while sitting and standing. In mild cases, some restless or shuffling or tramping movements are present, which the individual is able to easily suppress when asked to do so. In severe ‘4’ cases, these movements are continuous and difficult to control with effort, and patient is often unable to remain seated or standing for many minutes, without walking or pacing in that period.

#### Other

Dystonia is characterized by sustained muscle contractions resulting in twisting and repetitive movements or abnormal postures. Dystonia is difficult to rate as many different parts of the body are potentially affected, and its severity changes depending upon the posture and activity of the involved part. The overall rating takes into account the body regions affected, the severity of the movement and the duration for which it is present. For rating of minimal ‘1’, dystonia is present occasionally (e.g., grimace or moth movement or blinking or occasional pulling of neck muscle) or only during an action. In mild cases, the movement is mild or present for <50% of the time and does not cause impairment. In moderate cases, movement is more persistent, and some impairment is present. In severe cases, considerable disability and/or distress ensue, and the movement is present all of the time.

Myoclonus refers to brief, sudden, involuntary muscle jerks. Some jerks, such as ‘sleep starts’ and hiccups are common and normal. Sometimes jerks may develop in patients on psychotropic drugs, in particular antidepressants and lithium carbonate. They may occur spontaneously at rest, or in response to stimuli, and may be infrequent or occur every few seconds. Their rating will depend upon their frequency, severity (or amplitude) and stimulus-sensitivity. Minimal ‘1’ rating refers to occasional jerks or trace movements, and mild ‘2’ myoclonus occurs many times a minute and is easily visible. In severe ‘4’ cases, the jerks are of large amplitude and occur almost every second, disabling the patient.

#### Global ratings

The examiner rates the patient’s disability and distress due to the movements as an overall rating, taking all movements into consideration, but accounting for the disability or distress due to other factors, including the mental illness. Disability refers to the functional impairment produced by the movement disorder, whereas distress is subjective and may be due to social embarrassment, interference with function or pain. A particular movement, such as orolingual dyskinesia, may be very distressing but may not impair the patient’s functioning. On the other hand, severe dystonia can be both severely distressing and disabling.

## Notes

### Competing Interest Statement

PSS has received honorariums from Biogen Australia and Roche Australia as a member of their advisory committees.

### Funding Statement

This study did not receive any funding.

### Author Declarations

Human Research Ethics Committee of the South-Eastern Sydney Local Health District gave ethical approval for this work.

